# PEPFAR Spending Types and Reduction in HIV Infection Rates

**DOI:** 10.1101/2022.11.14.22282303

**Authors:** Stephen Walker

## Abstract

Since 2004, PEPFAR has invested over $100 billion in fighting HIV, primarily targeting sub-Saharan Africa. This study examines the effectiveness of spending types as defined by the program. In this study, I take the spending data published by PEPFAR that was categorized into key focus areas that include (1) Care & Treatment, (2) Testing, (3) Prevention, (4) Socioeconomic, and (5) Above-site, Program Management and Other, and estimate the effectiveness of these categories on a key outcome variable: new HIV infections. I also estimate the same regressions along two dimensions: (1) pre and post PEPFAR 2.0 that was implemented in 2014, and (2) the 12 sub-Saharan focus countries versus the non-sub-Saharan countries. Data was collected from public sources including PEPFAR, the GHO, and the World Bank and a total of $61.5 billion in spending was identified from 2005-2021 covering 54 target countries.

**Main Outcome(s) and Measure(s):** The marginal dollar spent on prevention activities experienced the highest incremental effect of reducing new HIV infections in targeted countries from 2005-2021. The coefficient on Prevention spending was -0.715 (t-stat of -2.83), which was highly significant at the 1 percent level. However, much of this effect was driven by pre-2014 spending before PEPFAR 2.0 was implemented. Post 2014, socioeconomic programs were measured to have the greatest marginal effect driven by non-sub-Saharan countries. Overall effectiveness of spending increased post PEPFAR 2.0 for non-Sub-Saharan countries but declined for sub-Saharan countries.

## Introduction

The President’s Emergency Plan for AIDS Relief (PEPFAR) was created in 2003 and annual funding for the program has grown from $2.2 billion in 2004 to 7.0 billion in 2022 (KFF 2022). PEPFAR was designed to address the problem that “more than 90% of all HIV infections were occurring in resource-limited countries, particularly in sub-Saharan Africa, where patients had little or no access to antiretroviral medications” (Fauci and Eisinger 2018). In addition to treatment, “PEPFAR has also provided some of the critical workforce, organizational, and physical infrastructure to address other concerns” (Fauci and Eisinger 2018).

Various studies have examined PEPFAR funding on health outcomes. Duber, et al. (2010) published a retrospective analysis and compared PEPFAR focus versus non-focus counties on various national health indicators (e.g., adult mortality and incidence of TB) and found that countries were either slightly worse off or not different. The authors concluded that PEPFAR “may have little or no impact on health outcomes not explicitly targeted.” Another paper by Chin, et al. (2015) applied a fixed-effects panel regression analysis on twelve African countries from 2002-2010 and found that HIV infection rates decreased by 0.355 percentage points for every 1 percentage point increase in funding per GDP. While the effect was statistically significant, they concluded that “the reduction rate should be higher.” This study extends this earlier work by Chin, et al. and explores the cross-sectional variation in total funding for PEPFAR countries. This study focuses on the subcategories of spending measured by PEPFAR that include care and treatment, testing, prevention, socio-economic spending, and other types of spending that include above-site preparation, program management, and pipeline spending. The purpose of this study is to document the effects for these spending categories that may help policy makers better allocate capital resources. Does prevention spending outperform socioeconomic spending at the margin or vice-versa? This study documents these differences that may be helpful to PEPFAR administrators when allocating spending budgets.

Data was collected from public sources. PEPFAR funding data comes from the PEPFAR website^1^. New HIV infection rates per thousand in population was sourced from the SDGHIV file from the GHO Odata api^2^. Population used to scale PEPFAR funding comes from the World Bank file.^3^

Table 1 summarizes the spending identified by PEPFAR by type and year. A total of $61.5 billion was identified from 2005-2021. Care & Treatment represents 46.1 percent of the spend. Above-site, program management and other spending consumed 29.1 percent of the budget. Testing was 7.2 percent, prevention was 9.5 percent, and socioeconomic was 8.2 percent. A total of 54 countries were identified in the data. Table 2 summarizes the data for the 12 sub-Saharan countries including Botswana, Cote d’Ivoire, Ethiopia, Kenya, Mozambique, Namibia, Nigeria, Rwanda, South Africa, Tanzania, Uganda, and Zambia where a total of $51.9 billion was identified representing 84 percent of the total dollars in the data. The remaining 42 non-sub-Saharan countries totaled $9.6 billion, which represents the remaining 16 percent of the spending.

**Table 1:** Summary Data by Year (All Countries) The Total dollars spent (in millions) and fraction of total spending as by identified PEPFAR category for years 2005-2021. A total of $61.5 billion was identified in the data.

| Year | Total Spending | Care & Treatment | Testing | Prevention | Socioeconomic | Above-Site, Program Management, Other |
| --- | --- | --- | --- | --- | --- | --- |
| *\$ in millions* | | | | | | |
| 2005 | \$ 1,069.5 | \$ 509.7 | \$ 96.4 | \$ 95.7 | \$ 95.2 | \$ 272.6 |
| 2006 | \$ 1,522.0 | \$ 796.1 | \$ 114.4 | \$ 124.7 | \$ 174.7 | \$ 312.1 |
| 2007 | \$ 2,556.8 | \$ 1,353.3 | \$ 223.2 | \$ 207.4 | \$ 289.5 | \$ 483.4 |
| 2008 | \$ 3,505.2 | \$ 1,786.9 | \$ 269.4 | \$ 295.0 | \$ 409.6 | \$ 744.2 |
| 2009 | \$ 3,553.4 | \$ 1,675.0 | \$ 265.1 | \$ 343.6 | \$ 387.8 | \$ 882.0 |
| 2010 | \$ 3,586.2 | \$ 1,686.5 | \$ 267.1 | \$ 350.4 | \$ 389.6 | \$ 892.5 |
| 2011 | \$ 3,693.6 | \$ 1,690.4 | \$ 336.6 | \$ 387.9 | \$ 398.3 | \$ 880.4 |
| 2012 | \$ 3,778.2 | \$ 1,662.4 | \$ 316.5 | \$ 425.1 | \$ 376.3 | \$ 997.8 |
| 2013 | \$ 3,191.5 | \$ 1,390.8 | \$ 272.6 | \$ 430.5 | \$ 266.7 | \$ 830.8 |
| 2014 | \$ 4,694.0 | \$ 1,541.9 | \$ 327.5 | \$ 391.2 | \$ 284.7 | \$ 2,148.8 |
| 2015 | \$ 3,979.8 | \$ 1,738.4 | \$ 253.6 | \$ 286.3 | \$ 234.8 | \$ 1,466.7 |
| 2016 | \$ 4,002.9 | \$ 2,062.0 | \$ 291.6 | \$ 302.2 | \$ 260.1 | \$ 1,087.0 |
| 2017 | \$ 3,990.5 | \$ 2,042.1 | \$ 238.2 | \$ 273.0 | \$ 254.8 | \$ 1,182.4 |
| 2018 | \$ 4,350.5 | \$ 2,040.2 | \$ 293.2 | \$ 338.9 | \$ 291.3 | \$ 1,386.8 |
| 2019 | \$ 4,539.5 | \$ 2,322.8 | \$ 286.0 | \$ 525.6 | \$ 325.3 | \$ 1,079.8 |
| 2020 | \$ 4,554.2 | \$ 1,778.0 | \$ 275.8 | \$ 454.9 | \$ 260.0 | \$ 1,785.6 |
| 2021 | \$ 4,909.5 | \$ 2,256.5 | \$ 278.3 | \$ 584.5 | \$ 321.6 | \$ 1,468.5 |
| **Total** | **\$ 61,477.3** | **\$ 28,333.2** | **\$ 4,405.4** | **\$ 5,816.8** | **\$ 5,020.6** | **\$ 17,901.4** |
| *Fraction of Total Spending* |  |  |  |  |  |  |
| 2005 | 100.0% | 47.7% | 9.0% | 8.9% | 8.9% | 25.5% |
| 2006 | 100.0% | 52.3% | 7.5% | 8.2% | 11.5% | 20.5% |
| 2007 | 100.0% | 52.9% | 8.7% | 8.1% | 11.3% | 18.9% |
| 2008 | 100.0% | 51.0% | 7.7% | 8.4% | 11.7% | 21.2% |
| 2009 | 100.0% | 47.1% | 7.5% | 9.7% | 10.9% | 24.8% |
| 2010 | 100.0% | 47.0% | 7.4% | 9.8% | 10.9% | 24.9% |
| 2011 | 100.0% | 45.8% | 9.1% | 10.5% | 10.8% | 23.8% |
| 2012 | 100.0% | 44.0% | 8.4% | 11.3% | 10.0% | 26.4% |
| 2013 | 100.0% | 43.6% | 8.5% | 13.5% | 8.4% | 26.0% |
| 2014 | 100.0% | 32.8% | 7.0% | 8.3% | 6.1% | 45.8% |
| 2015 | 100.0% | 43.7% | 6.4% | 7.2% | 5.9% | 36.9% |
| 2016 | 100.0% | 51.5% | 7.3% | 7.5% | 6.5% | 27.2% |
| 2017 | 100.0% | 51.2% | 6.0% | 6.8% | 6.4% | 29.6% |
| 2018 | 100.0% | 46.9% | 6.7% | 7.8% | 6.7% | 31.9% |
| 2019 | 100.0% | 51.2% | 6.3% | 11.6% | 7.2% | 23.8% |
| 2020 | 100.0% | 39.0% | 6.1% | 10.0% | 5.7% | 39.2% |
| 2021 | 100.0% | 46.0% | 5.7% | 11.9% | 6.6% | 29.9% |
| **Total** | **100.0%** | **46.1%** | **7.2%** | **9.5%** | **8.2%** | **29.1%** |

**Table 2:** Summary Data by Year (12 Sub-Saharan Countries) The Total dollars spent (in millions) and fraction of total spending as by identified PEPFAR category for years 2005-2021. A total of $51.9 billion was identified in the data, or 84 percent of the total dollars.

| Year | Total Spending | Care & Treatment | Testing | Prevention | Socioeconomic | Above-Site, Program Management, Other |
| --- | --- | --- | --- | --- | --- | --- |
| *\$ in millions* | | | | | | |
| 2005 | \$ 1,002.3 | \$ 476.5 | \$ 91.8 | \$ 87.3 | \$ 90.2 | \$ 256.6 |
| 2006 | \$ 1,370.4 | \$ 734.0 | \$ 100.6 | \$ 97.9 | \$ 157.9 | \$ 280.0 |
| 2007 | \$ 2,300.7 | \$ 1,245.0 | \$ 197.9 | \$ 159.7 | \$ 261.5 | \$ 436.7 |
| 2008 | \$ 3,195.6 | \$ 1,660.9 | \$ 239.6 | \$ 237.4 | \$ 382.2 | \$ 675.6 |
| 2009 | \$ 3,231.1 | \$ 1,573.0 | \$ 238.6 | \$ 289.0 | \$ 359.1 | \$ 771.5 |
| 2010 | \$ 3,231.1 | \$ 1,573.0 | \$ 238.6 | \$ 289.0 | \$ 359.1 | \$ 771.5 |
| 2011 | \$ 3,240.3 | \$ 1,548.6 | \$ 295.3 | \$ 307.4 | \$ 360.1 | \$ 729.0 |
| 2012 | \$ 3,227.3 | \$ 1,493.1 | \$ 269.8 | \$ 336.0 | \$ 335.8 | \$ 792.6 |
| 2013 | \$ 2,687.3 | \$ 1,226.2 | \$ 228.6 | \$ 334.3 | \$ 236.1 | \$ 662.2 |
| 2014 | \$ 3,858.8 | \$ 1,363.8 | \$ 272.9 | \$ 294.9 | \$ 250.7 | \$ 1,676.4 |
| 2015 | \$ 3,308.7 | \$ 1,476.1 | \$ 207.6 | \$ 216.8 | \$ 206.0 | \$ 1,202.3 |
| 2016 | \$ 3,352.7 | \$ 1,761.9 | \$ 249.3 | \$ 242.6 | \$ 229.6 | \$ 869.2 |
| 2017 | \$ 3,326.9 | \$ 1,711.1 | \$ 194.5 | \$ 225.5 | \$ 222.8 | \$ 973.1 |
| 2018 | \$ 3,599.0 | \$ 1,717.3 | \$ 242.0 | \$ 279.5 | \$ 238.9 | \$ 1,121.3 |
| 2019 | \$ 3,746.7 | \$ 1,935.8 | \$ 216.7 | \$ 417.6 | \$ 269.0 | \$ 907.6 |
| 2020 | \$ 3,517.1 | \$ 1,482.7 | \$ 173.1 | \$ 350.0 | \$ 209.2 | \$ 1,302.1 |
| 2021 | \$ 3,688.6 | \$ 1,800.8 | \$ 179.4 | \$ 444.3 | \$ 238.4 | \$ 1,025.7 |
| **Total** | **\$ 51,884.8** | **\$ 24,779.6** | **\$ 3,636.2** | **\$ 4,609.2** | **\$ 4,406.6** | **\$ 14,453.3** |
| *Fraction of Total Spending* |  |  |  |  |  |  |
| 2005 | 100.0% | 47.5% | 9.2% | 8.7% | 9.0% | 25.6% |
| 2006 | 100.0% | 53.6% | 7.3% | 7.1% | 11.5% | 20.4% |
| 2007 | 100.0% | 54.1% | 8.6% | 6.9% | 11.4% | 19.0% |
| 2008 | 100.0% | 52.0% | 7.5% | 7.4% | 12.0% | 21.1% |
| 2009 | 100.0% | 48.7% | 7.4% | 8.9% | 11.1% | 23.9% |
| 2010 | 100.0% | 48.7% | 7.4% | 8.9% | 11.1% | 23.9% |
| 2011 | 100.0% | 47.8% | 9.1% | 9.5% | 11.1% | 22.5% |
| 2012 | 100.0% | 46.3% | 8.4% | 10.4% | 10.4% | 24.6% |
| 2013 | 100.0% | 45.6% | 8.5% | 12.4% | 8.8% | 24.6% |
| 2014 | 100.0% | 35.3% | 7.1% | 7.6% | 6.5% | 43.4% |
| 2015 | 100.0% | 44.6% | 6.3% | 6.6% | 6.2% | 36.3% |
| 2016 | 100.0% | 52.6% | 7.4% | 7.2% | 6.8% | 25.9% |
| 2017 | 100.0% | 51.4% | 5.8% | 6.8% | 6.7% | 29.2% |
| 2018 | 100.0% | 47.7% | 6.7% | 7.8% | 6.6% | 31.2% |
| 2019 | 100.0% | 51.7% | 5.8% | 11.1% | 7.2% | 24.2% |
| 2020 | 100.0% | 42.2% | 4.9% | 10.0% | 5.9% | 37.0% |
| 2021 | 100.0% | 48.8% | 4.9% | 12.0% | 6.5% | 27.8% |
| **Total** | **100.0%** | **47.8%** | **7.0%** | **8.9%** | **8.5%** | **27.9%** |

## Methods

This paper measures the effect that each spending type has on new HIV infection rates. Six fixed effects panel regressions were estimated that take the following form:

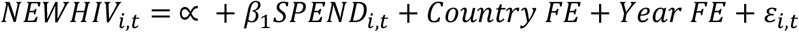

The dependent variable NEWHIV is defined as the new HIV infection rate per thousand in population. The SPEND variable is the total dollar value for each spending category identified by PEPFAR, scaled by population. Country and year fixed effects were included in each regression. The SPEND variable represents one of Total Spending, or each of the subcategories including Care & Treatment, Testing, Prevention, Socioeconomic, and above-site/program management/other types.

A final model is estimated that takes the following form, which includes all subcategories and all other that add up to Total Spending.

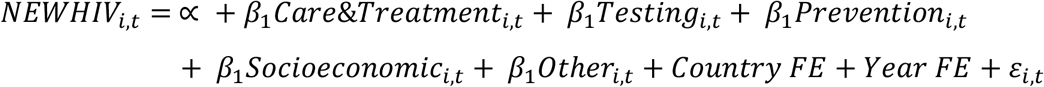

These models are estimated for the overall sample from 2005-2021, and for the period 2015-2021, which represents a new regime in the data under PEPFAR 2.0 (explained in detail in the discussion section). Additional regressions explore differences between sub-Saharan countries, which represent the bulk of dollars over this period, and non-sub-Saharan countries, where spending nearly doubled from 2015 levels.

Standard errors are clustered at the firm level. The theoretical prediction for the sign of each variable is negative as the goal is to decrease new HIV infections.

## Results & Discussion

The results of the regressions for the entire sample are summarized in Table 4. Consistent with the prior literature, the measured coefficient is negative on total PEPFAR funding and was significant at the one percent level (t-stat of -4.94). The coefficient of -0.119 can be interpreted in the following way. For every $1 per person increase in spending for a given country, the new HIV infection rate per 1,000 decreases by 0.119. Compared to the mean rate in the data of 2.5 per 1,000, this effect appears small, consistent with the previous literature.

**Table 3:** Summary Data by Year (42 Non-Sub-Saharan Countries) The Total dollars spent (in millions) and fraction of total spending as by identified PEPFAR category for years 2005-2021. A total of $9.6 billion was identified in the data, or 16 percent of the total dollars.

| Year | Total Spending | Care & Treatment | Testing | Prevention | Socioeconomic | Above-Site, Program Management, Other |
| --- | --- | --- | --- | --- | --- | --- |
| *\$ in millions* | | | | | | |
| 2005 | \$ 67.3 | \$ 33.2 | \$ 4.5 | \$ 8.4 | \$ 5.1 | \$ 16.0 |
| 2006 | \$ 151.6 | \$ 62.1 | \$ 13.7 | \$ 26.8 | \$ 16.8 | \$ 32.1 |
| 2007 | \$ 256.1 | \$ 108.2 | \$ 25.4 | \$ 47.7 | \$ 28.0 | \$ 46.7 |
| 2008 | \$ 309.6 | \$ 126.1 | \$ 29.8 | \$ 57.6 | \$ 27.5 | \$ 68.6 |
| 2009 | \$ 322.2 | \$ 102.0 | \$ 26.5 | \$ 54.6 | \$ 28.7 | \$ 110.5 |
| 2010 | \$ 355.0 | \$ 113.5 | \$ 28.5 | \$ 61.4 | \$ 30.6 | \$ 121.0 |
| 2011 | \$ 453.3 | \$ 141.8 | \$ 41.3 | \$ 80.5 | \$ 38.2 | \$ 151.5 |
| 2012 | \$ 550.9 | \$ 169.3 | \$ 46.7 | \$ 89.2 | \$ 40.5 | \$ 205.2 |
| 2013 | \$ 504.2 | \$ 164.7 | \$ 44.0 | \$ 96.2 | \$ 30.6 | \$ 168.6 |
| 2014 | \$ 835.2 | \$ 178.1 | \$ 54.6 | \$ 96.3 | \$ 33.9 | \$ 472.4 |
| 2015 | \$ 671.1 | \$ 262.3 | \$ 46.0 | \$ 69.5 | \$ 28.8 | \$ 264.5 |
| 2016 | \$ 650.3 | \$ 300.1 | \$ 42.3 | \$ 59.6 | \$ 30.5 | \$ 217.8 |
| 2017 | \$ 663.6 | \$ 331.1 | \$ 43.8 | \$ 47.5 | \$ 32.1 | \$ 209.2 |
| 2018 | \$ 751.4 | \$ 322.9 | \$ 51.2 | \$ 59.3 | \$ 52.4 | \$ 265.5 |
| 2019 | \$ 792.8 | \$ 387.1 | \$ 69.2 | \$ 107.9 | \$ 56.4 | \$ 172.2 |
| 2020 | \$ 1,037.1 | \$ 295.3 | \$ 102.6 | \$ 104.9 | \$ 50.8 | \$ 483.5 |
| 2021 | \$ 1,220.9 | \$ 455.8 | \$ 99.0 | \$ 140.2 | \$ 83.2 | \$ 442.8 |
| **Total** | **\$ 9,592.5** | **\$ 3,553.5** | **\$ 769.2** | **\$ 1,207.6** | **\$ 614.1** | **\$ 3,448.1** |
| *Fraction of Total Spending* |  |  |  |  |  |  |
| 2005 | 100.0% | 49.4% | 6.7% | 12.5% | 7.5% | 23.8% |
| 2006 | 100.0% | 41.0% | 9.1% | 17.7% | 11.1% | 21.2% |
| 2007 | 100.0% | 42.3% | 9.9% | 18.6% | 11.0% | 18.2% |
| 2008 | 100.0% | 40.7% | 9.6% | 18.6% | 8.9% | 22.2% |
| 2009 | 100.0% | 31.6% | 8.2% | 16.9% | 8.9% | 34.3% |
| 2010 | 100.0% | 32.0% | 8.0% | 17.3% | 8.6% | 34.1% |
| 2011 | 100.0% | 31.3% | 9.1% | 17.8% | 8.4% | 33.4% |
| 2012 | 100.0% | 30.7% | 8.5% | 16.2% | 7.4% | 37.2% |
| 2013 | 100.0% | 32.7% | 8.7% | 19.1% | 6.1% | 33.4% |
| 2014 | 100.0% | 21.3% | 6.5% | 11.5% | 4.1% | 56.6% |
| 2015 | 100.0% | 39.1% | 6.9% | 10.4% | 4.3% | 39.4% |
| 2016 | 100.0% | 46.1% | 6.5% | 9.2% | 4.7% | 33.5% |
| 2017 | 100.0% | 49.9% | 6.6% | 7.2% | 4.8% | 31.5% |
| 2018 | 100.0% | 43.0% | 6.8% | 7.9% | 7.0% | 35.3% |
| 2019 | 100.0% | 48.8% | 8.7% | 13.6% | 7.1% | 21.7% |
| 2020 | 100.0% | 28.5% | 9.9% | 10.1% | 4.9% | 46.6% |
| 2021 | 100.0% | 37.3% | 8.1% | 11.5% | 6.8% | 36.3% |
| **Total** | **100.0%** | **37.0%** | **8.0%** | **12.6%** | **6.4%** | **35.9%** |

**Table 4:** PEPFAR Spending and New HIV Infections (2005-2021) Dependent variable in each regression is New HIV Infections per 1,000 people sourced from the World Health Organization. PEPFAR dollar spending by category is sourced from PEPFAR data and is scaled by population. All models include year and country fixed effects. Robust t-statistics clustered by country are reported in parentheses (***p<0.01, ** p<0.05, * p<0.1)

|  | Total Funding | Care & Treatment | Testing | Prevention | Socioeconomic | Above-Site, Program Management, Other | All Spending Types Included |
| --- | --- | --- | --- | --- | --- | --- | --- |
| Total Spending | -0.119***<br>(-4.94) |  |  |  |  |  |  |
| Care & Treatment |  | -0.205***<br>(-3.28) |  |  |  |  | -0.239***<br>(-5.53) |
| Testing |  |  | -0.388<br>(-0.91) |  |  |  | 0.271<br>(1.60) |
| Prevention |  |  |  | -0.715***<br>(-2.83) |  |  | -0.639***<br>(-3.29) |
| Socioeconomic |  |  |  |  | -0.470<br>(-1.29) |  | 0.825***<br>(4.68) |
| Other |  |  |  |  |  | -0.205***<br>(-5.05) | -0.184***<br>(-5.76) |
| Constant | 4.306***<br>(10.72) | 4.430***<br>(8.43) | 4.945***<br>(8.59) | 4.653***<br>(14.03) | 4.864***<br>(13.74) | 4.851***<br>(15.23) | 5.104***<br>(20.88) |
| Observations | 489 | 488 | 486 | 470 | 406 | 404 | 404 |
| R-squared | 0.653 | 0.595 | 0.461 | 0.606 | 0.531 | 0.587 | 0.780 |
| Number of cc | 47 | 47 | 47 | 47 | 40 | 40 | 40 |
| Year FE | YES | YES | YES | YES | YES | YES | YES |
| Country FE | YES | YES | YES | YES | YES | YES | YES |

Exploring how each spending type matters, the second column from Table 4 shows the estimate for Care & Treatment spending, which was -0.205 (t-stat of -3.28) and significant at the 1 percent level, which is a near doubling of the overall effect measured (−0.119 with t-stat of - 4.94). Services included in this category include programs such as HIV clinical services, HIV laboratory services, HIV Drugs, and other care and treatment. Among the four key spending categories, it is measured as the most significant variable. The third column explores Testing spending, which includes facility-based testing, community-based testing, and other testing programs. The results documented here imply that there is no significance with increased testing spending though the size effect is larger than the total spending at -0.388. While testing is important to reduce new infections, there may be wide variation in outcomes from country to country which contributes to the lack of significance. The fourth column reports Prevention spending, which includes community mobilization, Prep, Condom and lubricant programming, medication assisted therapy, primary prevention for HIV and sexual violence programs. In Table 4, prevention spending shows the highest marginal effect among the spending types with a coefficient of -0.715 (t-stat of -2.83), which is 6x the effect of total overall spending. The fifth column reports Socioeconomic spending, which includes case management, economic strengthening, education assistance, food and nutrition, psychosocial support, legal, human right and protection, and other socioeconomic spending. Similar to Testing, Socioeconomic spending was not significant. The sixth column captures above-site, program management and other unidentified spending. Results are similar to Care & Treatment in this model and significant at the 1 percent level. Lastly, all variables that sum to total funding were estimated in the seventh column. Controlling for each of the other spending types, Care & Treatment is the most statistically significant among the four key categories (t-stat of -5.53), but Prevention spending measured the highest with an estimated coefficient of -0.639 (t-stat of -3.29).

Banigbe, et al. (2019) studied the effect of PEPFAR policy change in Nigeria with the introduction of PEPFAR 2.0 that occurred in 2014. “The transition to PEPFAR 2.0 with its focus on country ownership was accompanied by substantial funding cuts” and “Funding cutbacks have been associated with compromised quality of care” (Banigbe, et al. 2019). This discontinuity is observed in the data for 2014 in Table 1. Therefore, to explore whether PEPFAR 2.0 had any effect on these measures, Table 5 Panel A re-estimates the previous regressions, but for 2015-2021. Table 5 Panel B repeats this process for 2005-2013. Both avoid the 2014 transition year in the estimation. The results from the panels in Table 5 show that, while prevention spending still matters, Socioeconomic programs were measured to have the largest marginal effect post 2014 with high statistical significance (coef. Of -0.944; t-stat of -7.43). The overall effect of total spending improves slightly from -0.093 (t-stat -6.48) in Panel B to -0.140 (t-stat -5.71) in Panel A.

**Table 5:**
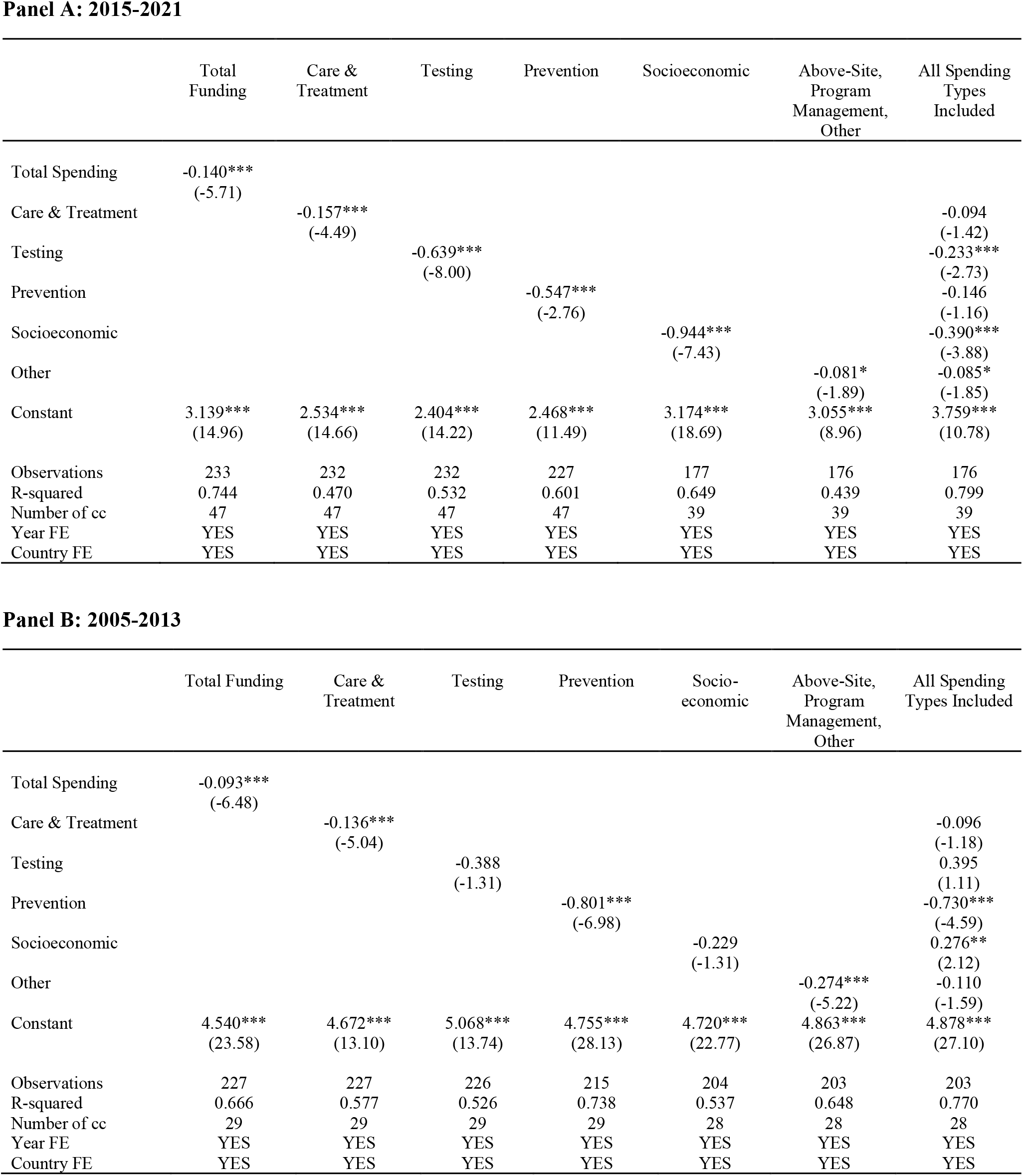
PEPFAR Spending and New HIV Infections. Dependent variable in each regression is New HIV Infections per 1,000 people sourced from the World Health Organization. PEPFAR dollar spending by category is sourced from PEPFAR data and is scaled by population. All models include year and country fixed effects. Robust t-statistics clustered by country are reported in parentheses (***p<0.01, ** p<0.05, * p<0.1)

**Panel A: 2015-2021**
|  | Total<br>Funding | Care &<br>Treatment | Testing | Prevention | Socioeconomic | Above-Site,<br>Program<br>Management,<br>Other | All Spending<br>Types<br>Included |
| --- | --- | --- | --- | --- | --- | --- | --- |
| Total Spending | -0.140***<br>(-5.71) |  |  |  |  |  |  |
| Care & Treatment |  | -0.157***<br>(-4.49) |  |  |  |  | -0.094<br>(-1.42) |
| Testing |  |  | -0.639***<br>(-8.00) |  |  |  | -0.233***<br>(-2.73) |
| Prevention |  |  |  | -0.547***<br>(-2.76) |  |  | -0.146<br>(-1.16) |
| Socioeconomic |  |  |  |  | -0.944***<br>(-7.43) |  | -0.390***<br>(-3.88) |
| Other |  |  |  |  |  | -0.081*<br>(-1.89) | -0.085*<br>(-1.85) |
| Constant | 3.139***<br>(14.96) | 2.534***<br>(14.66) | 2.404***<br>(14.22) | 2.468***<br>(11.49) | 3.174***<br>(18.69) | 3.055***<br>(8.96) | 3.759***<br>(10.78) |
| Observations | 233 | 232 | 232 | 227 | 177 | 176 | 176 |
| R-squared | 0.744 | 0.470 | 0.532 | 0.601 | 0.649 | 0.439 | 0.799 |
| Number of cc | 47 | 47 | 47 | 47 | 39 | 39 | 39 |
| Year FE | YES | YES | YES | YES | YES | YES | YES |
| Country FE | YES | YES | YES | YES | YES | YES | YES |

**Panel B: 2005-2013**
|  | Total Funding | Care &<br>Treatment | Testing | Prevention | Socio-<br>economic | Above-Site,<br>Program<br>Management,<br>Other | All Spending<br>Types Included |
| --- | --- | --- | --- | --- | --- | --- | --- |
| Total Spending | -0.093***<br>(-6.48) |  |  |  |  |  |  |
| Care & Treatment |  | -0.136***<br>(-5.04) |  |  |  |  | -0.096<br>(-1.18) |
| Testing |  |  | -0.388<br>(-1.31) |  |  |  | 0.395<br>(1.11) |
| Prevention |  |  |  | -0.801***<br>(-6.98) |  |  | -0.730***<br>(-4.59) |
| Socioeconomic |  |  |  |  | -0.229<br>(-1.31) |  | 0.276**<br>(2.12) |
| Other |  |  |  |  |  | -0.274***<br>(-5.22) | -0.110<br>(-1.59) |
| Constant | 4.540***<br>(23.58) | 4.672***<br>(13.10) | 5.068***<br>(13.74) | 4.755***<br>(28.13) | 4.720***<br>(22.77) | 4.863***<br>(26.87) | 4.878***<br>(27.10) |
| Observations | 227 | 227 | 226 | 215 | 204 | 203 | 203 |
| R-squared | 0.666 | 0.577 | 0.526 | 0.738 | 0.537 | 0.648 | 0.770 |
| Number of cc | 29 | 29 | 29 | 29 | 28 | 28 | 28 |
| Year FE | YES | YES | YES | YES | YES | YES | YES |
| Country FE | YES | YES | YES | YES | YES | YES | YES |

The bulk of PEPFAR dollars are spent in 12 sub-Saharan countries, though growth in new spending has occurred in countries outside this region (Tables 2 and 3). Therefore, it is worth exploring whether there are differences along this dimension. Table 6 reports the results for the 12 sub-Saharan countries only. Based on these results, overall effectiveness declined, and other effects were attenuated, particularly Prevention. Socio-economic programs and testing programs gained significance, however. Table 7 reports the same results, but for the non-Sub-Saharan countries. Results improved from -0.110 in the pre-2014 period to -0.173 in the PEPFAR 2.0 period. Individual effects of each of the subcategories were similar, except for socio-economic activities which improved substantially. Overall, Socio-economic programs experienced the highest marginal effect outside the sub-Saharan sphere while prevention programs matter the most in sub-Saharan countries.

**Table 6:** PEPFAR Spending and New HIV Infections Sub-saharan Only. Dependent variable in each regression is New HIV Infections per 1,000 people sourced from the World Health Organization. PEPFAR dollar spending by category is sourced from PEPFAR data and is scaled by population. All models include year and country fixed effects. Robust t-statistics clustered by country are reported in parentheses (***p<0.01, ** p<0.05, * p<0.1)

**Panel A: 2015-2021**
|  | Total Funding | Care & Treatment | Testing | Prevention | Socio-economic | Above-Site, Program Management, Other | All Spending Types Included |
| --- | --- | --- | --- | --- | --- | --- | --- |
| Total Spending | -0.050*<br>(-1.82) |  |  |  |  |  |  |
| Care & Treatment |  | -0.079**<br>(-2.52) |  |  |  |  | -0.056<br>(-0.65) |
| Testing |  |  | -0.269***<br>(-3.51) |  |  |  | 0.019<br>(0.10) |
| Prevention |  |  |  | -0.223**<br>(-2.93) |  |  | -0.267**<br>(-3.02) |
| Socioeconomic |  |  |  |  | -0.278**<br>(-2.92) |  | 0.517*<br>(1.90) |
| Other |  |  |  |  |  | 0.033**<br>(2.69) | -0.009<br>(-0.25) |
| Constant | 3.234***<br>(11.37) | 2.940***<br>(16.80) | 2.766***<br>(20.56) | 2.782***<br>(18.36) | 2.818***<br>(24.66) | 2.426***<br>(14.13) | 2.735***<br>(7.91) |
| Observations | 77 | 77 | 77 | 77 | 77 | 77 | 77 |
| R-squared | 0.640 | 0.659 | 0.618 | 0.693 | 0.602 | 0.601 | 0.726 |
| Number of cc | 11 | 11 | 11 | 11 | 11 | 11 | 11 |
| Year FE | YES | YES | YES | YES | YES | YES | YES |
| Country FE | YES | YES | YES | YES | YES | YES | YES |

**Panel B: 2005-2013**
|  | Total Funding | Care & Treatment | Testing | Prevention | Socio-economic | Above-Site, Program Management, Other | All Spending Types Included |
| --- | --- | --- | --- | --- | --- | --- | --- |
| Total Spending | -0.097***<br>(-4.39) |  |  |  |  |  |  |
| Care & Treatment |  | -0.132***<br>(-5.45) |  |  |  |  | -0.094<br>(-1.21) |
| Testing |  |  | -0.241<br>(-0.63) |  |  |  | 0.414<br>(1.10) |
| Prevention |  |  |  | -0.710***<br>(-4.51) |  |  | -0.580**<br>(-2.97) |
| Socioeconomic |  |  |  |  | -0.068<br>(-0.41) |  | 0.220*<br>(1.90) |
| Other |  |  |  |  |  | -0.296***<br>(-4.56) | -0.136<br>(-1.34) |
| Constant | 5.508***<br>(27.78) | 5.488***<br>(17.52) | 5.635***<br>(18.68) | 5.577***<br>(23.40) | 5.107***<br>(23.01) | 5.531***<br>(34.22) | 5.461***<br>(23.58) |
| Observations | 108 | 108 | 108 | 99 | 99 | 99 | 99 |
| R-squared | 0.789 | 0.689 | 0.636 | 0.845 | 0.669 | 0.805 | 0.879 |
| Number of cc | 11 | 11 | 11 | 11 | 11 | 11 | 11 |
| Year FE | YES | YES | YES | YES | YES | YES | YES |
| Country FE | YES | YES | YES | YES | YES | YES | YES |

**Table 7:** PEPFAR Spending and New HIV Infections 2015-2021 – Non-sub-saharan Only. Dependent variable in each regression is New HIV Infections per 1,000 people sourced from the World Health Organization. PEPFAR dollar spending by category is sourced from PEPFAR data and is scaled by population. All models include year and country fixed effects. Robust t-statistics clustered by country are reported in parentheses (***p<0.01, ** p<0.05, * p<0.1)

**Panel A: 2015-2021**
|  | Total Funding | Care & Treatment | Testing | Prevention | Socio-economic | Above-Site, Program Management, Other | All Spending Types Included |
| --- | --- | --- | --- | --- | --- | --- | --- |
| Total Spending | -0.173***<br>(-12.01) |  |  |  |  |  |  |
| Care & Treatment |  | -0.211***<br>(-11.73) |  |  |  |  | -0.176***<br>(-7.11) |
| Testing |  |  | -0.774***<br>(-19.03) |  |  |  | -0.097<br>(-1.18) |
| Prevention |  |  |  | -0.946***<br>(-5.50) |  |  | -0.163<br>(-1.34) |
| Socioeconomic |  |  |  |  | -1.105***<br>(-15.74) |  | -0.395**<br>(-2.65) |
| Other |  |  |  |  |  | -0.158***<br>(-3.91) | -0.166***<br>(-12.07) |
| Constant | 2.657***<br>(17.93) | 2.256***<br>(13.59) | 2.149***<br>(9.02) | 2.390***<br>(13.05) | 3.172***<br>(15.22) | 3.165***<br>(8.11) | 3.880***<br>(25.78) |
| Observations | 156 | 155 | 155 | 150 | 100 | 99 | 99 |
| R-squared | 0.882 | 0.457 | 0.564 | 0.742 | 0.707 | 0.496 | 0.930 |
| Number of cc | 36 | 36 | 36 | 36 | 28 | 28 | 28 |
| Year FE | YES | YES | YES | YES | YES | YES | YES |
| Country FE | YES | YES | YES | YES | YES | YES | YES |

**Panel B: 2005-2013**
|  | Total Funding | Care & Treatment | Testing | Prevention | Socio-economic | Above-Site, Program Management, Other | All Spending Types Included |
| --- | --- | --- | --- | --- | --- | --- | --- |
| Total Spending | -0.110***<br>(-4.43) |  |  |  |  |  |  |
| Care & Treatment |  | -0.217***<br>(-4.84) |  |  |  |  | -0.077<br>(-0.46) |
| Testing |  |  | -0.839***<br>(-3.13) |  |  |  | 0.126<br>(0.21) |
| Prevention |  |  |  | -0.981***<br>(-5.70) |  |  | -1.147***<br>(-5.02) |
| Socioeconomic |  |  |  |  | -0.833***<br>(-3.62) |  | 0.960<br>(1.60) |
| Other |  |  |  |  |  | -0.259**<br>(-2.88) | -0.225<br>(-1.58) |
| Constant | 2.663***<br>(17.85) | 2.698***<br>(20.53) | 3.359***<br>(88.20) | 3.634***<br>(32.85) | 3.562***<br>(26.99) | 3.545***<br>(21.58) | 4.010***<br>(20.12) |
| Observations | 119 | 119 | 118 | 116 | 105 | 104 | 104 |
| R-squared | 0.521 | 0.469 | 0.416 | 0.636 | 0.505 | 0.478 | 0.669 |
| Number of cc | 18 | 18 | 18 | 18 | 17 | 17 | 17 |
| Year FE | YES | YES | YES | YES | YES | YES | YES |
| Country FE | YES | YES | YES | YES | YES | YES | YES |

## Conclusion

This study documents the cross-sectional effects of subcategories of PEPFAR spending from 2005-2021 along two dimensions including the pre and post PEPFAR 2.0 period that occurred in 2014, and sub-Saharan focus countries versus those outside this region. One key finding is that prevention spending has the largest effect on new HIV infection rates over the other spending categories, overall. For sub-Saharan countries from 2015-2021, prevention spending was shown to have the highest marginal effect when considering all spending types. For non-sub-Saharan countries, socioeconomic programs were shown to be most effective on the key outcome measure of reducing new HIV infections.

## Data Availability

All data produced in the present study are available upon reasonable request to the authors

## Footnotes

1 https://data.pepfar.gov/

2 https://www.who.int/data/gho/info/gho-odata-api

3 https://data.worldbank.org/indicator/SP.POP.TOTL

## References

Banigbe, B., Audet, C. M., Okonkwo, P., Arije, O. O., Bassi, E., Clouse, K., … & Ahonkhai, A. A. (2019). Effect of PEPFAR funding policy change on HIV service delivery in a large HIV care and treatment network in Nigeria. PLoS One, 14(9), e0221809.

Chin, R. J., Sangmanee, D., & Piergallini, L. (2015). PEPFAR funding and reduction in HIV infection rates in 12 focus sub-Saharan African countries: a quantitative analysis. International Journal of MCH and AIDS, 3(2), 150.

El-Sadr, W. M., Holmes, C. B., Mugyenyi, P., Thirumurthy, H., Ellerbrock, T., Ferris, R., … & Whiteside, A. (2012). Scale-up of HIV treatment through PEPFAR: a historic public health achievement. Journal of acquired immune deficiency syndromes (1999), 60(Suppl 3), S96.

Fauci, A. S., & Eisinger, R. W. (2018). PEPFAR—15 years and counting the lives saved. New England Journal of Medicine, 378(4), 314–316.

Goosby, E., Von Zinkernagel, D., Holmes, C., Haroz, D., & Walsh, T. (2012). Raising the bar: PEPFAR and new paradigms for global health. JAIDS Journal of Acquired Immune Deficiency Syndromes, 60, S158–S162.

Katz, I. T., Bassett, I. V., & Wright, A. A. (2013). PEPFAR in transition—implications for HIV care in South Africa. New England Journal of Medicine, 369(15), 1385–1387.

Kim, Y. (2022). The effectiveness of the US President’s Emergency Plan for AIDS Relief (PEPFAR) in responding to HIV/AIDS in Four African Countries. The International Journal of Health Planning and Management.

Lee, M. M., & Izama, M. P. (2015). Aid externalities: evidence from PEPFAR in Africa. World Development, 67, 281–294.

Menzies, N. A., Berruti, A. A., Berzon, R., Filler, S., Ferris, R., Ellerbrock, T. V., & Blandford, J. M. (2011). The cost of providing comprehensive HIV treatment in PEPFAR-supported programs. AIDS (London, England), 25(14), 1753.

Padian, N. S., Holmes, C. B., McCoy, S. I., Lyerla, R., Bouey, P. D., & Goosby, E. P. (2011). Implementation science for the US President’s Emergency Plan for AIDS Relief (PEPFAR). JAIDS Journal of Acquired Immune Deficiency Syndromes, 56(3), 199–203.

